# Morning SARS-CoV-2 testing yields better detection of infection due to higher viral loads in saliva and nasal swabs upon waking

**DOI:** 10.1101/2022.03.02.22271724

**Authors:** Alexander Viloria Winnett, Michael K. Porter, Anna E. Romano, Emily S. Savela, Reid Akana, Natasha Shelby, Jessica A. Reyes, Noah W. Schlenker, Matthew M. Cooper, Alyssa M. Carter, Jenny Ji, Jacob T. Barlow, Colten Tognazzini, Matthew Feaster, Ying-Ying Goh, Rustem F. Ismagilov

**Affiliations:** California Institute of Technology, 1200 E. California Blvd., Pasadena, CA, USA 91125; City of Pasadena Public Health Department, 1845 N. Fair Oaks Ave., Pasadena, CA, USA 91103

**Keywords:** COVID-19, SARS-CoV-2, pandemic, diagnostic, testing, screening, sensitivity, saliva, nasal swab, outpatient, ambulatory, mild, circadian, rhythm, morning, evening, collection, sample, viral load

## Abstract

**Background:** The analytical sensitivities of SARS-CoV-2 diagnostic tests span 6 orders of magnitude. Optimizing sample-collection methods to achieve the most reliable detection for a given sensitivity would increase the effectiveness of testing and minimize COVID-19 outbreaks.

**Methods:** From September 2020 to April 2021 we performed a household-transmission study in which participants self-collected samples every morning and evening throughout acute SARS-CoV-2 infection. Seventy mildly symptomatic participants collected saliva and, of those, 29 also collected nasal-swab samples. Viral load was quantified in 1194 saliva and 661 nasal-swab samples using a high-analytical-sensitivity RT-qPCR assay (LOD, 1,000 SARS-CoV-2 RNA copies/mL).

**Findings:** Viral loads in both saliva and nasal-swab samples were significantly higher in morning-collected samples than evening-collected samples after symptom onset. We used these quantitative measurements to infer which diagnostic tests would have detected infection (based on sample type and test analytical sensitivity). We find that morning collection would have resulted in significantly improved detection and that this advantage would be most pronounced for tests with low to moderate analytical sensitivity, which would likely have missed infections if sampling in the evening.

**Interpretation:** Collecting samples for COVID-19 testing in the morning offers a simple and low-cost improvement to clinical diagnostic sensitivity of low- to moderate-analytical-sensitivity tests. The phenomenon of higher viral loads in the morning may also have implications related to when transmission is more likely to occur.

**Funding:** Bill & Melinda Gates Foundation, Ronald and Maxine Linde Center for New Initiatives (Caltech), Jacobs Institute for Molecular Engineering for Medicine (Caltech)

**RESEARCH IN CONTEXT:** *Evidence before this study:* Reliable COVID-19 diagnostic testing is critical to reducing transmission of SARS-CoV-2 and reducing cases of severe or fatal disease, particularly in areas with limited vaccine access or uptake. Saliva and anterior-nares nasal swabs are common sample types; however, different diagnostic tests using these sample types have a range of analytical sensitivities spanning 6 orders of magnitude, with limits of detection (LODs) between 10^2^ and 10^8^ genomic copy equivalents of SARS-CoV-2 RNA (copies) per mL of sample. Due to limitations in clinical laboratory capacity, many low-resource settings rely on COVID-19 tests that fall on the moderate (LODs of 10^4^ to 10^5^ copies/mL) to lower (LODs of 10^5^ to 10^8^ copies/mL) end of this spectrum of analytical sensitivity. Alterations in sample collection methods, including time of sample collection, may improve the performance of these diagnostics.

*Added value of this study:* This study quantifies viral loads from saliva and nasal-swab samples that were longitudinally self-collected by symptomatic patients in the morning immediately after waking and in the evening just prior to sleeping throughout the course of acute SARS-CoV-2 infection. The study cohort was composed of mildly or moderately symptomatic individuals (outpatients). This analysis demonstrates significantly higher viral loads in samples collected in the morning, relative to those collected in the evening. When using moderate to lower analytical sensitivity test methods, these loads are inferred to result in significantly better detection of infected individuals in the morning.

*Implications of available evidence:* These findings suggest that samples collected in the morning immediately after waking will better detect SARS-CoV-2 infection in symptomatic individuals tested by moderate to lower analytical sensitivity COVID-19 diagnostic tests (LODs at or above 10^4^ viral copies per mL of sample), such as many rapid antigen tests currently available.

## INTRODUCTION

Although vaccination has substantially reduced hospitalizations and death from COVID-19, limited vaccine uptake and availability and the potential for breakthrough infections (particularly with novel viral variants) support the continued necessity for diagnostic testing and subsequent isolation of infected individuals.^1,2^ Optimizing how diagnostics are used can enhance our ability to combat the COVID-19 pandemic.

Nasopharyngeal swab, anterior-nares swab, mid-turbinate swab, oropharyngeal swab, buccal swab, gingival crevicular fluid, sputum, tracheal aspirate, and saliva have all been put forth and compared as diagnostic specimens for the detection of SARS-CoV-2 infection. Work done by many groups,^3-5^ including ours^6^, has suggested that SARS-CoV-2 is detectable, albeit at low viral loads, in saliva before anterior nares nasal-swab samples. However, conflicting results have been reported in head-to-head comparisons of saliva to other specimen types in cross-sectional studies.

The lack of clarity on which sample type is most reliable for SARS-CoV-2 detection is likely due to the dynamic nature of viral load in different sample types through the course of an infection^3,6-11^ and the differences in analytical sensitivity of diagnostic assays used in the comparisons. Currently available SARS-CoV-2 diagnostics span a very wide (6 orders of magnitude) range of analytical sensitivities; for example, the RT-PCR PerkinElmer New Coronavirus Nucleic Acid Detection Kit (LOD of 180 NDU/mL)^12^ to Coris BioConcept rapid antigen lateral flow assay COVID-19 Ag Respi-Strip (LOD approximately 4×10^7^ copies/mL).^13^ Tests with moderate analytical sensitivity (LOD of 10^4^ to 10^5^ copies/mL of sample) or low analytical sensitivity (LOD of 10^5^ to 10^8^ copies/mL of sample) are being increasingly used, particularly for at-home and rapid screening testing, and in areas of the world with limited laboratory capacity.^14-16^

In addition, how samples are collected can affect detectability of SARS-CoV-2 in the sample. Because SARS-CoV-2, like other pathogens, may exhibit circadian rhythms to replication kinetics,^17,18^ we hypothesized that collection time may impact SARS-CoV-2 viral load in clinical respiratory specimens, and therefore detectability of initial infection. Simple, low-cost changes to sample collection protocols that significantly improve the clinical sensitivity of COVID-19 diagnostics offer an immediately actionable opportunity to improve testing with existing diagnostics, which would be particularly valuable in settings with limited laboratory capacity that rely on tests with low analytical sensitivity.

We conducted a COVID-19 household transmission study^9,19^ where participants prospectively self-collected saliva and nasal-swab samples twice per day (in the morning, and in the evening). From mildly symptomatic participants, we compared SARS-CoV-2 viral loads in morning- and evening-collected samples to determine if a pattern to the time of day could be leveraged to improve detection and diagnosis of SARS-CoV-2 infection.

## METHODS

### Study Design

Participants were recruited for participation in a COVID-19 household-transmission study as previously described.^9,19^ This study was reviewed and approved by the Institutional Review Board of the California Institute of Technology (protocol #20-1026). All adult participants provided written informed consent, and minors (ages 6-17) provided assent accompanied by parental permission. Household members aged 6 years and above were eligible, which involved a questionnaire with demographic/health information and COVID-19 infection history, and self-collection of respiratory samples every morning and every night (described below).

For participants who were SARS-CoV-2-positive when initially enrolled in the study, symptom onset was defined as the date of first symptom reported in the questionnaire. For participants who entered the study SARS-CoV-2-negative but had unrelated symptoms, symptom onset was the first instance of a new COVID-19-like symptom or an increase in symptom severity following their first SARS-CoV-2-positive sample.

### Sample Collection

Participants self-collected anterior nares nasal-swab and saliva samples in the Spectrum SDNA-1000 Saliva Collection Kit (Spectrum Solutions LLC, Draper, UT, USA), at home twice per day (after waking up and before going to bed), per manufacturer’s guidelines (although Spectrum devices are not currently authorized for the collection of nasal-swab samples). One participant self-collected both anterior nares nasal-swab and saliva samples in Nest Viral Transport Media (VTM) [Cat. NST-NST-202117, Stellar Scientific, Baltimore, MD, USA], and three collected only nasal-swab samples in VTM. Participants were instructed not to ingest anything, smoke, or brush their teeth for at least 30 minutes prior to collection. For nasal-swab collection, participants were asked to gently blow their noses before swabbing (four complete rotations with gentle pressure in each nostril) with one of three types of sterile flocked swabs: Nest Oropharyngeal Specimen Collection Swabs [Cat. NST-202003, Stellar Scientific, Baltimore, MD, USA], or Puritan HydraFlock Swab [Cat. 25-3000-H E30, Puritan, Guilford, ME, USA] or Copan USA FLOQSwab [Cat. 520CS01, VWR International, Radnor, PA, USA]. A parent/guardian assisted minors with collection. Immediately after collection, participants recorded the date and time and any symptoms experienced in the previous 12 hours. Samples collected between 4am and 12pm were defined as morning samples; samples collected between 3pm and 3am were defined as evening samples (**Figure S1**).

### Cohort of Individuals with SARS-CoV-2 Infection

We quantified saliva viral loads in samples from 72 SARS-CoV-2-infected individuals from 39 households in southern California between September 2020 and April 2021. Of these, 2 (2.7%) never reported experiencing symptoms, and were not included in subsequent analyses where results were binned relative to symptom onset. Of the 70 symptomatic individuals from 37 households included in the analyses (**Table 1**), 58 were positive and 12 were negative on enrollment in both sample types but became positive while enrolled in the study. Of the 58 cases positive on enrollment, 50 (86.2%) were already experiencing mild COVID-19 like symptoms and 8 (13.8%) were pre-symptomatic. Of the 20 individuals who were negative (12) and pre-symptomatic (8) on enrollment, COVID-19 symptoms first developed an average of 1.21 days after first positive test (either while enrolled, or via CLIA testing just prior to enrollment). Among the 14 adults (≥18 years old) this interval was on average, 0.92 days, whereas in the 6 children (6-17 years old), it was 1.81 days.

**Table 1.**
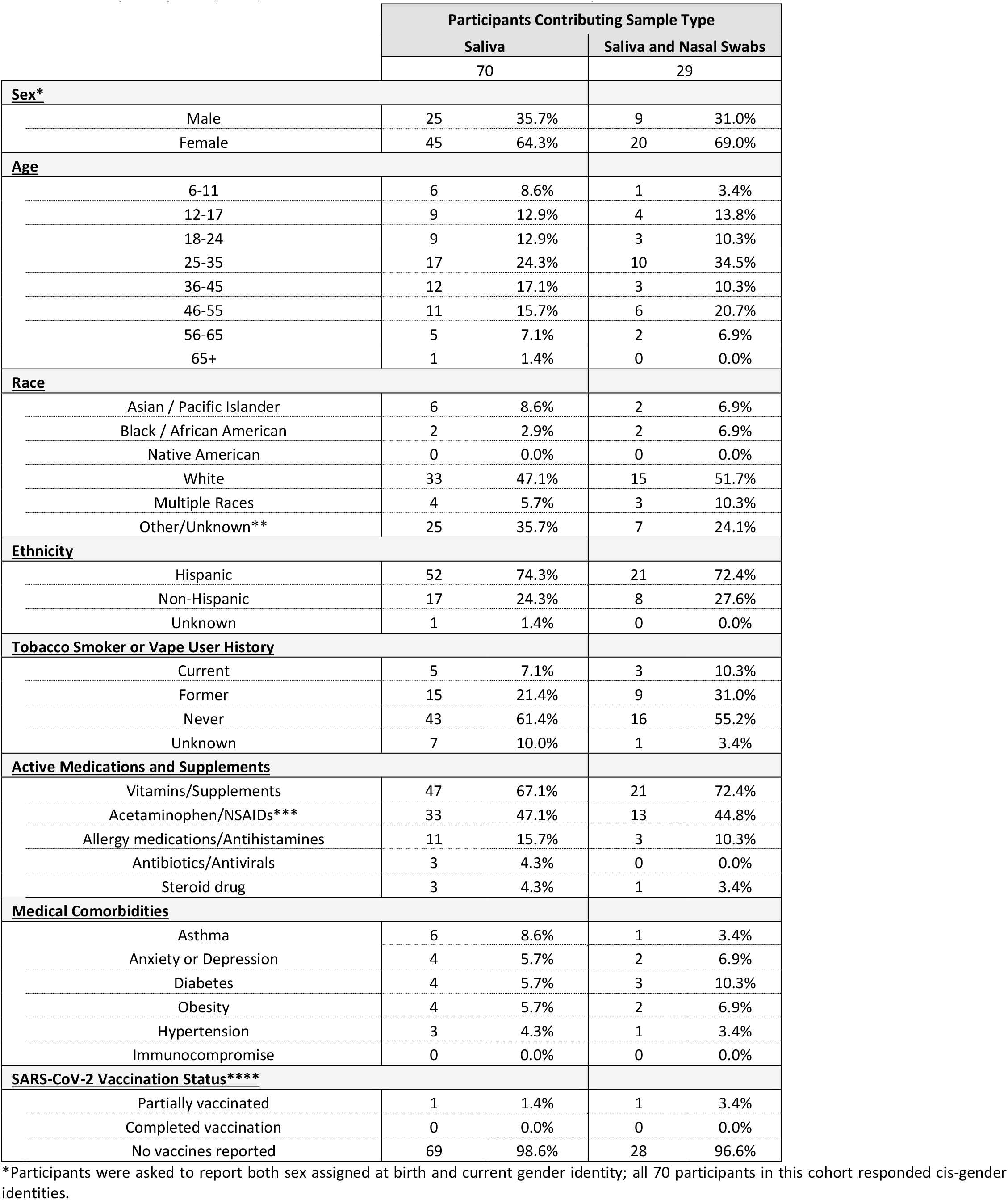
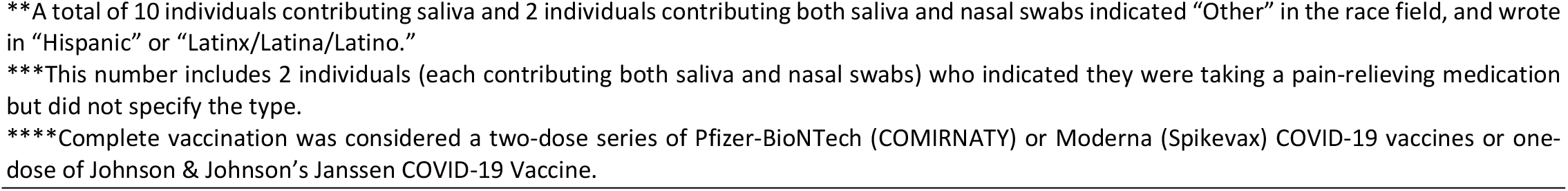
Demographics of Cohort Testing Positive for SARS-CoV-2. Data were collected via online questionnaire upon study enrollment. All participants (N=70) collected saliva; of these 70, 29 additionally collected nasal swabs.

|  |  | Participants Contributing Sample Type |  |  |  |
| --- | --- | --- | --- | --- | --- |
|  |  | Saliva |  | Saliva and Nasal Swabs |  |
|  |  | 70 |  | 29 |  |
| Sex* |  |  |  |  |  |
| Male |  | 25 | 35.7% | 9 | 31.0% |
| Female |  | 45 | 64.3% | 20 | 69.0% |
| Age |  |  |  |  |  |
| 6-11 |  | 6 | 8.6% | 1 | 3.4% |
| 12-17 |  | 9 | 12.9% | 4 | 13.8% |
| 18-24 |  | 9 | 12.9% | 3 | 10.3% |
| 25-35 |  | 17 | 24.3% | 10 | 34.5% |
| 36-45 |  | 12 | 17.1% | 3 | 10.3% |
| 46-55 |  | 11 | 15.7% | 6 | 20.7% |
| 56-65 |  | 5 | 7.1% | 2 | 6.9% |
| 65+ |  | 1 | 1.4% | 0 | 0.0% |
| Race |  |  |  |  |  |
| Asian / Pacific Islander |  | 6 | 8.6% | 2 | 6.9% |
| Black / African American |  | 2 | 2.9% | 2 | 6.9% |
| Native American |  | 0 | 0.0% | 0 | 0.0% |
| White |  | 33 | 47.1% | 15 | 51.7% |
| Multiple Races |  | 4 | 5.7% | 3 | 10.3% |
| Other/Unknown** |  | 25 | 35.7% | 7 | 24.1% |
| Ethnicity |  |  |  |  |  |
| Hispanic |  | 52 | 74.3% | 21 | 72.4% |
| Non-Hispanic |  | 17 | 24.3% | 8 | 27.6% |
| Unknown |  | 1 | 1.4% | 0 | 0.0% |
| Tobacco Smoker or Vape User History |  |  |  |  |  |
| Current |  | 5 | 7.1% | 3 | 10.3% |
| Former |  | 15 | 21.4% | 9 | 31.0% |
| Never |  | 43 | 61.4% | 16 | 55.2% |
| Unknown |  | 7 | 10.0% | 1 | 3.4% |
| Active Medications and Supplements |  |  |  |  |  |
| Vitamins/Supplements |  | 47 | 67.1% | 21 | 72.4% |
| Acetaminophen/NSAIDs*** |  | 33 | 47.1% | 13 | 44.8% |
| Allergy medications/Antihistamines |  | 11 | 15.7% | 3 | 10.3% |
| Antibiotics/Antivirals |  | 3 | 4.3% | 0 | 0.0% |
| Steroid drug |  | 3 | 4.3% | 1 | 3.4% |
| Medical Comorbidities |  |  |  |  |  |
| Asthma |  | 6 | 8.6% | 1 | 3.4% |
| Anxiety or Depression |  | 4 | 5.7% | 2 | 6.9% |
| Diabetes |  | 4 | 5.7% | 3 | 10.3% |
| Obesity |  | 4 | 5.7% | 2 | 6.9% |
| Hypertension |  | 3 | 4.3% | 1 | 3.4% |
| Immunocompromise |  | 0 | 0.0% | 0 | 0.0% |
| SARS-CoV-2 Vaccination Status**** |  |  |  |  |  |
| Partially vaccinated |  | 1 | 1.4% | 1 | 3.4% |
| Completed vaccination |  | 0 | 0.0% | 0 | 0.0% |
| No vaccines reported |  | 69 | 98.6% | 28 | 96.6% |

\*Participants were asked to report both sex assigned at birth and current gender identity; all 70 participants in this cohort responded cis-gender identities.
\*\*A total of 10 individuals contributing saliva and 2 individuals contributing both saliva and nasal swabs indicated “Other” in the race field, and wrote in “Hispanic” or “Latinx/Latina/Latino.”
\*\*\*This number includes 2 individuals (each contributing both saliva and nasal swabs) who indicated they were taking a pain-relieving medication but did not specify the type.
\*\*\*\*Complete vaccination was considered a two-dose series of Pfizer-BioNTech (COMIRNATY) or Moderna (Spikevax) COVID-19 vaccines or one-dose of Johnson & Johnson’s Janssen COVID-19 Vaccine.

From within the cohort of 70 individuals, 29 individuals from 16 households collected both saliva and nasal-swab sample types. Of these 29 individuals, 20 were positive in both sample types on enrollment, 2 were positive in saliva but initially negative in nasal swab, and 7 were initially negative in both sample types but became positive in both sample types; the viral loads and symptoms of these 7 individuals has been previously reported. ^6^ Of the 22 individuals positive in at least one sample type upon enrollment, 20 (90.1%) developed symptoms prior to enrollment and 2 (9.9%) were pre-symptomatic. No participants required hospitalization.

The mean age of the saliva cohort was 32.8 years (SD ±16.0 years), and 33.9 years (SD ±15.2 years) among those collecting both saliva and nasal swabs. Of 70 individuals providing saliva samples, 58 reported good, very good, or excellent health status. While enrolled, a total of 47 participants were taking vitamins and/or supplements, 33 were taking acetaminophen or an NSAID, 11 participants were taking an antihistamine and/or allergy medication, 2 participants were taking a statin (**Fig S2O, Fig S2U**), 2 participants were taking both an antibiotic and anti-seizure medication (**Fig S2AR, Fig S2AN**), and 3 individuals were taking a steroid drug (2 taking the inhaled medication fluticasone [**Fig S2N, Fig S2BP**], and 1 taking prednisone [**Fig S2BS**]). One participant (**Fig S2BT**) with Hepatitis C and HIV infection was taking Biktarvy (bictegravir/emtricitabine/tenofovir alafenamide). Additionally, one participant was pregnant (**Fig S2AB**). At the time of these participants’ enrollment in the study (September 2020 to April 2021), vaccines were either not available or limited to priority groups. Only one individual (**Fig S2H, Fig S3H**) reported receiving a COVID-19 vaccine (a first dose of the Pfizer-BioNTech COVID-19 vaccine, approximately three weeks prior to enrollment).

### Sample Extraction and Quantification of Viral Load by RT-qPCR

Sample processing was performed as previously described.^9^ Briefly, 400 μL of fluid from each saliva or nasal-swab sample was extracted using the MagMAX Viral/Pathogen Nucleic Acid Isolation Kit (ThermoFisher Scientific, Cat. A42352), followed by the CDC 2019-Novel Coronavirus (2019-nCoV) Real-Time RT-PCR Diagnostic Panel, which targets the *SARS-CoV-2 N1* and *N2* genes, as well as a human *RNase P* control. Our observed limit of detection (LOD) for this workflow, validated by spiking heat-inactivated SARS-CoV-2 particles (BEI Resources NR-52286) into human sample matrix (Lee Bio 991-05-P-PreC for saliva, and Lee Bio 991-13-P-PreC nasal fluid), was 1,000 genomic copy equivalents of SARS-CoV-2 RNA copies/mL of sample.^6^ For SARS-CoV-2 positive samples, the *N1* Ct value was converted to viral load using an equation derived from a standard curve of heat-inactivated SARS-CoV-2 particles spiked into human sample matrix validated by independent RT-ddPCR measurement.^6^ Of note, many COVID-19 diagnostics utilize *N* gene targets and *N* gene viral loads have been shown to track with other gene targets,^20^ suggesting that *N* gene quantification to viral load conversion would be representative to demonstrate a general phenomenon and relevant for diagnostics that detect other viral gene targets.

### Statistical analyses

Initial processing was performed in Python v3.8.2, where log transformed averages were calculated (**Fig1A, Fig1C**). Data was exported and differences in Ct from sequential samples and the average difference in Ct values was performed in Microsoft Excel (**Fig 2B, Fig 2C**). Plots were prepared in GraphPad Prism 9.2.0, including calculation of medians. For comparison of the differences between morning and evening viral loads and differences in Ct values, Mann-Whitney test was performed using GraphPad (**Fig 2**). One-sided binomial tests to compare inferred percentage of infections detectable by assays with varying LODs for samples collected in the morning or evening (**Fig 3**) was performed in Python v3.8.2 using the scipy.stats package.^21^

**Figure 1.**
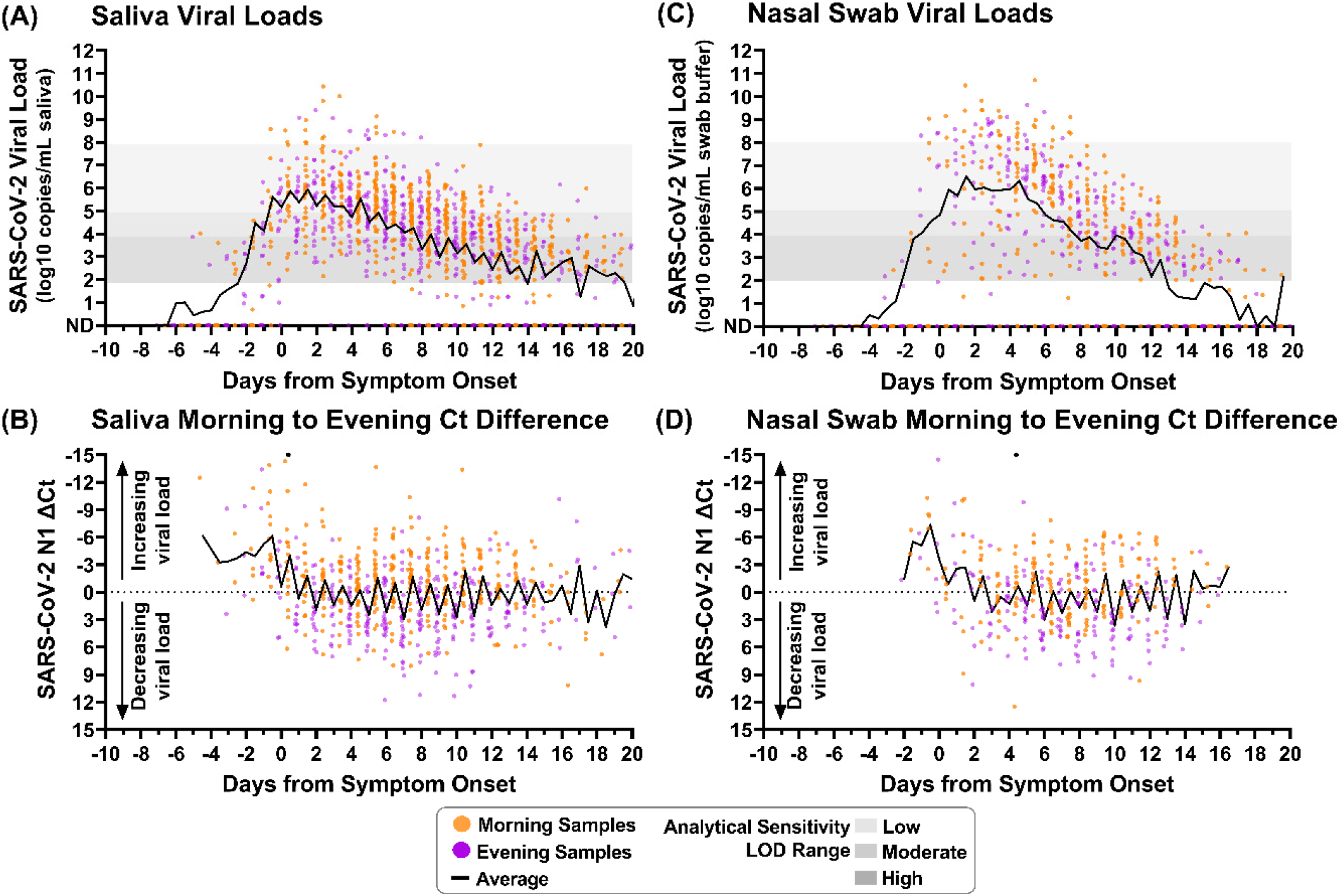
Saliva and nasal-swab samples collected in the morning and evening through the course of infection demonstrate differences in SARS-CoV-2 viral load. Black lines on each plot indicate the average viral load for each daily morning or evening sample collection window. Two black circles indicate morning sample measurements with Ct differences less than -15. **(A)** Saliva sample viral load (SARS-CoV-2 *N1* copies/mL saliva) as measured by RT-qPCR is plotted relative to symptom onset for 1194 samples. **(B)** The difference between morning and evening saliva *N1* Ct values is plotted relative to symptom onset for 703 sequential saliva samples. **(C)** Nasal-swab sample viral load (*N1* copies/mL saliva) as measured by RT-qPCR is plotted relative to symptom onset for 661 samples. **(D)** The difference between morning and evening nasal-swab *N1* Ct values is plotted relative to symptom onset for 385 sequential nasal-swab samples. Samples were designated as morning (orange) if collected between 4am and 12pm or evening (purple) if collected between 3pm and 3am. ND indicates Not Detected. Additional sample details provided in SI.

**Figure 2.**
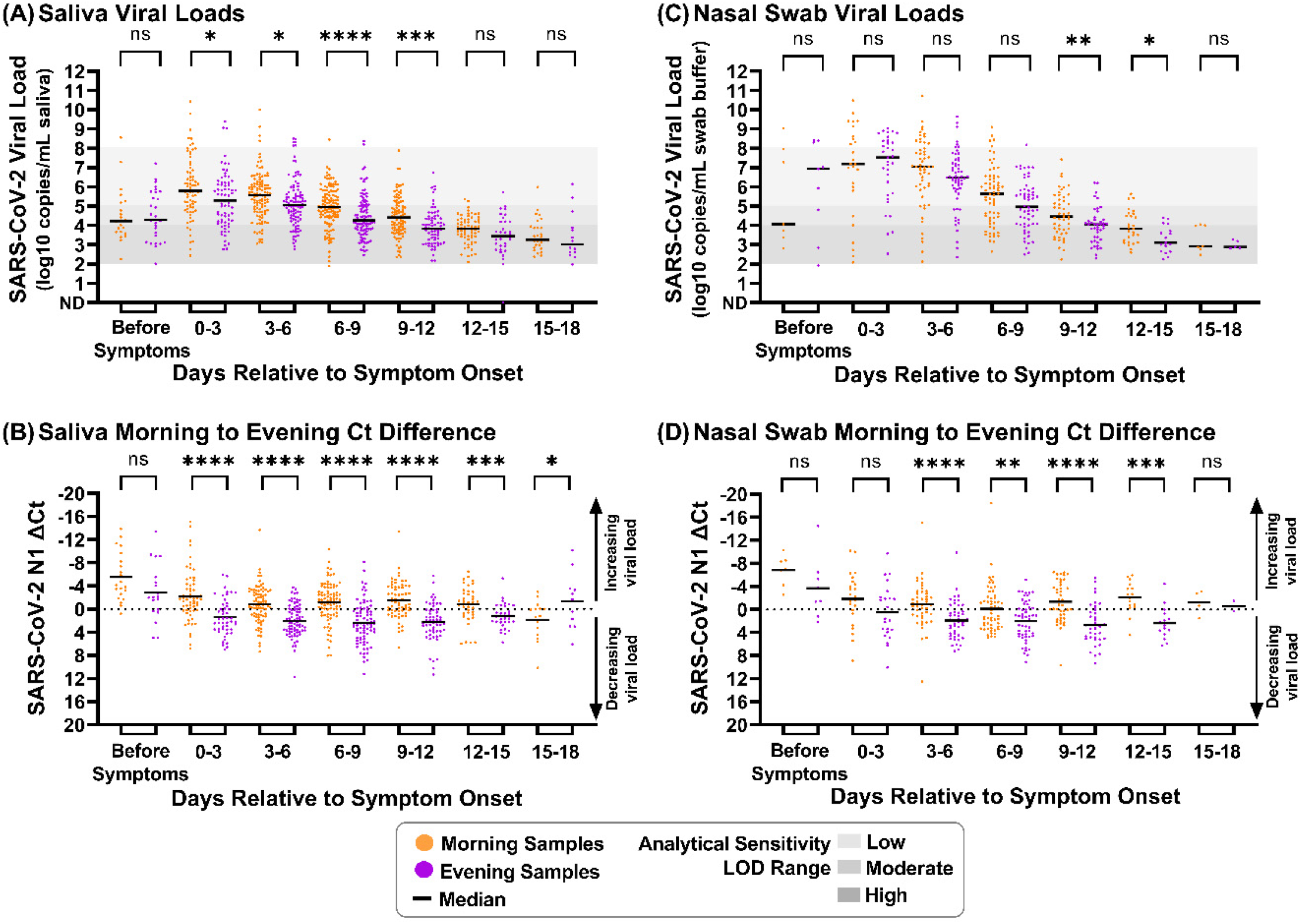
Morning viral loads are significantly higher than evening viral loads during most of SARS-CoV-2 infection. **(A)** Salivary SARS-CoV-2 *N1* viral loads are grouped by positive samples taken in the morning and evening, during different periods of infection. **(B)** Salivary SARS-CoV-2 *N1* changes in Ct value (ΔCt) are grouped by positive samples taken in the morning and evening, during different periods of infection. **(C)** Nasal-swab SARS-CoV-2 *N1* viral load are grouped by positive samples taken in the morning and evening, during different periods of infection. **(D)** Nasal-swab SARS-CoV-2 *N1* ΔCt are grouped by positive samples taken in the morning and evening, during different periods of infection. Comparison of viral loads by Mann-Whitney test: ns indicates nonsignificant, * indicates *P* < 0.05, ** indicates *P* < 0.01, **** indicates *P* < 0.001. Black lines indicate median viral load. ND indicates Not Detected.

**Figure 3.**
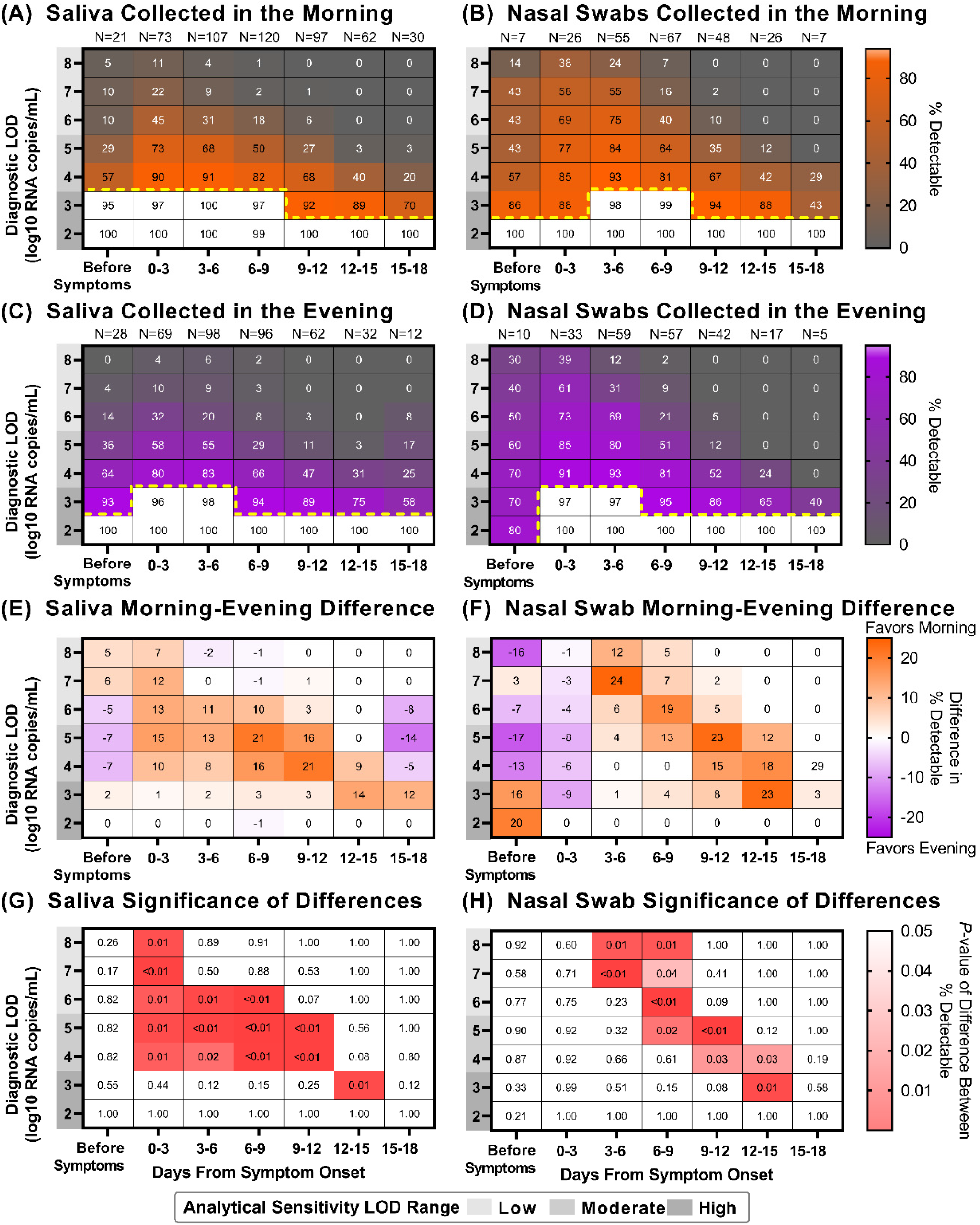
Saliva and Nasal-swab sample collection in the morning versus evening can yield different COVID-19 testing results depending on the limit of detection of the assay being used and the stage of infection. Heat maps showing the percentage of samples that would be considered positive when using a COVID-19 test with a given Limit of Detection (LOD), depicted on the y-axis, and based on the period of infection relative to symptom onset, depicted on the x-axis: **(A)** saliva collected in the morning **(B)** nasal swabs collected in the morning **(C)** saliva collected in the evening **(D)** nasal swabs collected in the evening. Yellow dotted lines indicate where sensitivity falls below 95%. For saliva **(E)** and nasal-swab **(F)** samples, a heat map showing the difference in percentage positivity between morning and evening samples from the same window of infection and test analytical sensitivities. For saliva **(G)** and nasal-swab **(H)** samples, a heat map of *P*-values resulting from a one-tailed binomial test that demonstrate the test sensitivities and periods of infection when morning nasal-swab samples yield a significantly higher percentage of samples inferred to be detectable for a given LOD and period of infection.

## RESULTS

### Timing of Morning and Evening Sample Collection

Viral load was quantified in a total of 1194 saliva samples and 661 nasal-swab samples. Across all samples, the distribution of sample collection times was roughly bimodal; we defined sampling upon waking (between 4am and 12pm) as morning, and before bed (between 3pm to 3am) as evening (**Figure S1**). For saliva samples, most participants (92%) had an average morning sample collection time between 7am and 10am. Evening sampling was slightly more variable; most participants (74%) had an average sample collection time between 8pm and 11pm. Each participant’s sampling time varied throughout the course of enrollment, though morning collection time was more consistent than evening; 76% of all participants collected at least 90% of morning samples within 90 minutes of their average morning sample collection time, compared to 37% of all participants collecting at least 90% of evening samples within 90 minutes of their average evening sample collection time. However, 81% of participants collected the majority (>50%) of evening samples within 90 minutes of their average evening sample collection time.

### Saliva and Nasal-Swab Samples Exhibit Higher Viral Loads in Morning Samples than Evening Samples Across the Course of Acute, Symptomatic Illness

Saliva and nasal-swab viral load profiles from most individuals (**Figure S2, Figure S3**) revealed a pattern of higher viral loads in samples collected in the morning compared with those collected in the evening. In samples from some individuals (e.g., **Figure S2A, Figure S3E**), fluctuations in both SARS-CoV-2 and human *RNase P* markers were observed, whereas in others *RNase P* remained stable and the pattern in SARS-CoV-2 viral load appeared to be independent of the host marker (e.g., **Figure S2AH, Figure S3N**).

Although direct comparison between all positive morning or evening samples demonstrates greater target abundance for both SARS-CoV-2 *N1* (**Figure S4A, Figure S4C**) and human *RNase P* (**Figure S4B, Figure S4D**), this comparison would be skewed by participants who contributed more samples and biased by sampling at different stages of the infection. To minimize these potential biases, the time of each sample collection was aligned relative to the date of symptom onset for that participant before plotting viral loads (**Figure S2, Figure S3**) and the average of log-transformed viral load values (**Figure 1A, Figure 1C**) for all saliva and nasal-swab samples at each timepoint. The average salivary viral load during each collection timepoint visually suggest higher viral loads in samples collected in the morning than in the evening. In both saliva and nasal-swab samples, the average viral load for 9 days after symptom onset falls within the reported range of LODs for moderate- (10^4^ to 10^5^ copies/mL) and low-analytical-sensitivity tests (10^5^ to 10^8^ copies/mL).

The pattern is further demonstrated when comparing the saliva *N1* Ct values between subsequent time points (**Figure 1B, Figure 1D**). RT-qPCR Ct values only for consecutively collected morning and evening samples were used to calculate the difference; negative or indeterminate samples were only used if directly proceeded by a positive sample collected in the pre-symptomatic phase of infection. A negative difference in Ct values indicates that viral load was increasing relative to the previous measurement, whereas a positive difference indicates that viral load was decreasing. Starting from symptom onset (Day 0), saliva samples collected in the morning typically exhibit a negative difference in Ct values relative to their preceding evening samples, whereas evening samples consistently have a positive difference. This indicates that throughout the course of symptomatic infection, morning samples typically result in relatively lower Ct values than evening samples. Although the nasal-swab viral loads do not exhibit as apparent of a pattern (**Figure 1C**), there are several points in the course of the infection (further described below) when this phenomenon is observed (**Figure 1D**).

To further illustrate the pattern observed in viral loads and changes in Ct values, samples were binned by symptom status: prior to symptom onset, and in 3-day intervals relative to the onset of symptoms. The 3-day interval was selected to capture reasonable resolution for infection stage, while also providing sufficient measurements to observe potential differences. Significantly higher morning viral loads were not observed prior to symptom onset in either sample type in the limited number of samples collected during this period. However, significantly higher viral loads (*P*<0.05, Mann-Whitney test) were observed in saliva samples collected in the morning for the first 12 days of symptomatic infection (**Figure 2A**), and differences in Ct values were significantly lower for morning samples in the 18 days following symptom onset (**Figure 2B**). Differences in Ct values were significantly lower (*P*<0.05, Mann-Whitney test) in morning nasal-swab samples from day 3 to day 15 of symptomatic infection (**Figure 2D**). However, nasal-swab samples collected in the morning did not have a statistically higher viral load compared to samples collected in the evening until 9 through 15 days after symptom onset (**Figure 2C**). Of note, nasal-swab viral load appears to increase more quickly to peak than salivary viral load (**Figure 1A, Figure 1C**), and nasal swabs also achieve higher peak viral loads (**Figure S4C**) than saliva (**Figure S4A**). The high rate of increase in viral load in nasal swabs likely obscures subtle daily fluctuations that are more apparent in saliva, where viral load rises more gradually. Nasal swabs appear to also be subject to more sampling variability (**Figure S4D, Figure S3**) than saliva (**Figure S4B, Figure S2**), evidenced by *RNase P* control marker Ct values.

### Saliva and Nasal-Swab Viral Loads in the Range of Moderate and Low Sensitivity Tests Underscore Utility of Morning Sampling

To understand the impact of higher viral loads in morning samples than evening samples, we used these quantitative viral load measurements to infer the performance of SARS-CoV-2 diagnostics with varying analytical sensitivities (i.e. LODs) at each collection time, for each sample type. The number of samples with viral load quantifications at or above a given LOD, out of the total number of samples for that time bin, was used to calculate the percentage of samples inferred to be reliably detected by a test with a given analytical sensitivity (**Figure 3**).

We hypothesized that sampling in the morning upon waking could detect significantly more infected individuals than sampling in the evening before bed. While analysis of the pre-symptomatic phase of infection is limited by few samples available for comparison, samples collected in the morning (**Figure 3A, B**) are more likely to be detected than those collected in the evening (**Figure 3C, D**) after symptom onset for both saliva and nasal-swab samples.

The differences in inferred detection between morning and evening collected saliva samples depend on the analytical sensitivity of the assay being used. When a high-analytical-sensitivity assay (LOD of 10^3^ copies/mL) is used, we infer non-significant differences in the detection of saliva (**Figure 3E**,**G**). However, an assay with a LOD of 10^6^ copies/mL was inferred to have significantly greater detection (up to 13%) of morning saliva samples for the first nine days of symptoms. The increase in detectable morning samples was statistically significant for diagnostic tests with LODs between 10^4^ to 10^8^ copies/mL, for the first twelve days of symptomatic infection.

Nasal-swab samples also exhibited differences in inferred detection between morning and evening collection that varied by analytical sensitivity. The inferred improvement was non-significant when using a high-analytical-sensitivity assay (LOD of 10^3^ copies/mL) up to 12 days from symptom onset, but larger and significant for moderate- to low-analytical-sensitivity assays. Significantly improved detection was observed for LODs 10^7^ to 10^8^ copies/mL from three to nine days after symptom onset, LODs 10^5^ to 10^8^ copies/mL from six to twelve days after symptom onset, and LODs 10^4^ to 10^5^ copies/mL from nine to fifteen days after symptom onset (**Figure 3F**,**H**).

These results suggest that collecting saliva or nasal-swab samples for SARS-CoV-2 diagnostic testing in the morning, immediately after waking up, can significantly improve detection of infected individuals particularly when only moderate- (LOD of 10^4^ to 10^5^ copies/mL of sample) to low- (LOD of 10^5^ to 10^8^ copies/mL of sample) analytical-sensitivity tests are available.

## DISCUSSION

In this study, we quantitatively measured SARS-CoV-2 viral load with high frequency (twice per day) longitudinally through the course of mild COVID-19 infection in saliva for 70 individuals, and in nasal swabs for 29 individuals. From these measurements, we identified a pattern of higher viral loads in saliva and nasal-swab samples collected in the morning after waking, compared to those collected in the evening. Similar observations have been reported for nasopharyngeal swabs,^22^ early morning versus spot oropharyngeal samples,^23^ early morning saliva versus nasopharyngeal swabs,^24^ as well as in wastewater surveillance.^25^

The reason that SARS-CoV-2 viral loads might be higher in the morning remains unknown. Similar to the improved performance of at-home pregnancy tests with morning urine due to accumulation of human chorionic gonadotropin,^26^ improved detection of SARS-CoV-2 may be the result of physical accumulation of material (e.g. cells, virions, nucleic acids) in the upper respiratory tract due to supine positioning (aiding mucociliary clearance) and/or the decreased rate of swallowing at night^27^. Higher morning viral loads due to physical accumulation of nucleic acids is supported by an increased abundance of the constitutively human *RNase P* target in saliva and nasal swabs samples collected in the morning (**Figure S2A, Figure S3A**). Human salivary production decreases overnight^28^ suggesting that higher morning viral loads could be due to a concentration of virus when saliva volume is lower.

Given that some individuals exhibit this phenomenon independent of human *RNase P* target abundance (**Figure S2B, Figure S3B**), a circadian rhythm in viral replication may also contribute. Regulation and responsiveness of the immune system has been linked to circadian rhythms,^29,30^ shown to affect SARS-CoV-2 infection of monocytes in cell culture^31^, and proposed as a modulating factor for COVID-19 severity and management.^32^ Others have proposed cellular interactions between viral proteins and circadian-dependent host signals,^33^ and demonstrated circadian-dependent entry and proliferation of SARS-CoV-2 in lung epithelial cell types in culture.^34^ Regardless of mechanism, because higher viral loads are associated with replication-competent culturable virus,^35,36^ these findings may also suggest a higher risk of transmission in the morning.

As many individuals remain unvaccinated and new variants emerge, it remains critical to identify infections, prompt isolation, trace and quarantine contacts, and initiate early treatment to improve efficacy. Much of the world lacks access to tests with high analytical sensitivity.^37-39^ Our findings suggest that strategically collecting samples in the morning immediately after waking up may improve the performance of available low- to moderate-analytical-sensitivity tests. Morning sampling will not raise the performance of tests with low analytical sensitivity to the levels of those with higher analytical sensitivity; however, even marginal improvements in detection have been shown to reduce deaths from COVID-19.^40^

This study is subject to four main limitations. First, we had a limited number of samples collected prior to the onset of symptoms, limiting our ability to discern a difference in detectability with morning or evening samples during the pre-symptomatic phase of infection. Second, this study was performed prior to the dominance of the Delta and Omicron variants of SARS-CoV-2, which may exhibit different viral-load kinetics. Third, host factors, including vaccination status, may also influence viral-load kinetics, but this study is not powered to demonstrate differences in viral-load profiles by demographic factors - data from all SARS-CoV-2-positive participants was aggregated for analysis. Fourth, this analysis involves inferring positivity by assays with varying analytical sensitivities (LODs), based on the quantitatively measured viral loads. A direct comparison with a specific test is needed to test real-world efficacy.

## Data Availability

All data produced are available online at CaltechDATA

https://data.caltech.edu/records/20049

## ACKNOWLEDGEMENTS

We thank Lauriane Quenee, Grace Fisher-Adams, Junie Hildebrandt, Megan Hyashi, RuthAnne Bevier, Chantal D’Apuzzo, Ralph Adolphs, Victor Rivera, Steve Chapman, Gary Waters, Leonard Edwards, Gaylene Ursua, Cynthia Ramos, and Shannon Yamashita for their assistance and advice on study implementation and/or administration. We thank Jessica Leong, Jessica Slagle, Mika Walton, Angel Navarro, Daniel Brenner, Ojas Pradhan, Si Hyung Jin and Mary Arrastia for volunteering their time to help with this study. We thank Angie Cheng, Susan Magdaleno, Christian Kis, Monica Herrera, and Zaina Lemeir for technical discussions regarding saliva extraction and detection. We thank Jennifer Fulcher, Debika Bhattacharya and Matthew Bidwell Goetz for their ideas on potential study populations and early study design. We thank Omai Garner and David Beenhouwer for providing materials for initial nasal-swab validation. We thank Martin Hill, Alma Sanchez, Scott Kim, Debbie Noble, Nina Paddock, Whitney Harrison, Emily Holman, Isaac Turner, Vivek Desai, Luke Wade, Tom Mayell, Stu Miller, Jennifer Howes, and Nari Shin for their support with recruitment. We thank Allison Rhines, Karen Heichman, and Dan Wattendorf for valuable discussions and guidance. We thank David Prober discussion of circadian rhythms and feedback on the manuscript. Finally, we thank all the case investigators and contact tracers at the Pasadena Public Health Department, the City of Long Beach Department of Health & Human Services and Caltech Student Wellness Services for their efforts in study recruitment and their work in the pandemic response.

## DATA AVAILABILITY

Data are available on CaltechDATA at: https://data.caltech.edu/records/20049

## SUPPLEMENTAL INFORMATION

## ADDITIONAL SAMPLE DETAILS FROM FIGURE 1

Viral load was quantified from an average of 32 saliva samples (SD ±6 samples) each from the 12 participants in the negative-on-enrollment cohort, while on average 13 saliva samples (SD ±10 samples) each were processed from 58 participants positive-on-enrollment (**Figure S2**). For nasal swabs, an average of 35 samples (SD ±7 samples) were quantified from 7 participants in the negative-on-enrollment cohort, while viral load was quantified in an average of 17 nasal-swab samples (SD ±9 samples) from 22 participants who were positive-on-enrollment (**Figure S3**).

**Figure S1.**
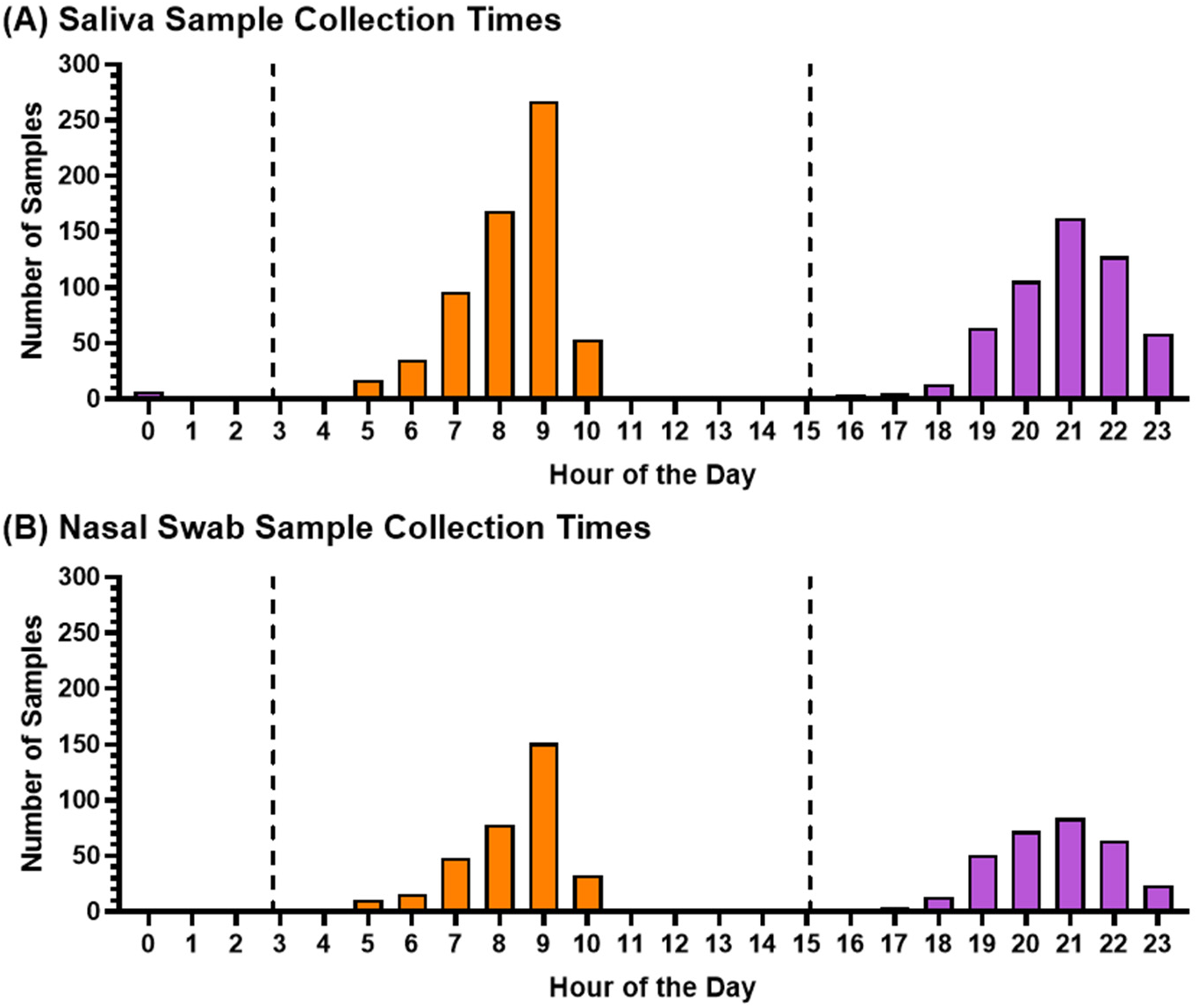
Frequency of Saliva and Nasal-Swab Sample Collection Times. Study participants either collected saliva only, or saliva then anterior nasal swab at the same time point, and were instructed to collect samples immediately after waking up and immediately before bed (see Methods for detailed instructions). The frequency of samples collected by each hour of the day is plotted for 1194 saliva samples **(A)** and 661 nasal-swab samples **(B)**. Dashed vertical line indicates cutoff for morning (3 AM to 12 PM) and evening (3 PM to 3 AM) collected samples used in this study.

**Figure S2.**
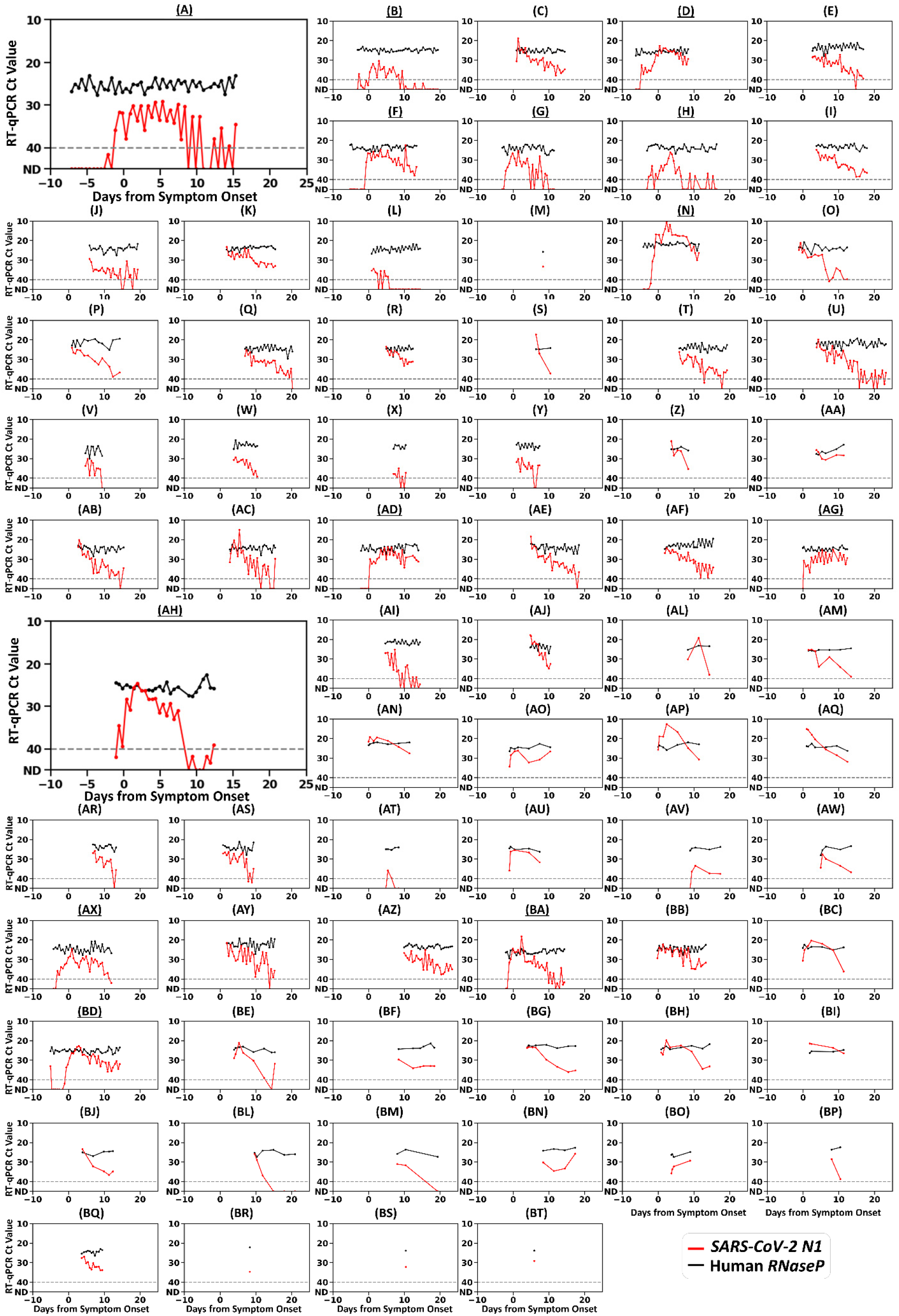
Individual salivary RT-PCR Ct measurements, for SARS-CoV-2 *N1* gene target (red) and human *RNase P* control gene target (black), relative to symptom onset. Matching panel labels correspond to the same participant shown in Figure S3. Underlined panel labels indicate that the participant converted from SARS-CoV-2-negative to -positive while enrolled in the study. Grey dashed line indicates Ct threshold for positivity. ND indicates Not Detected.

**Figure S3.**
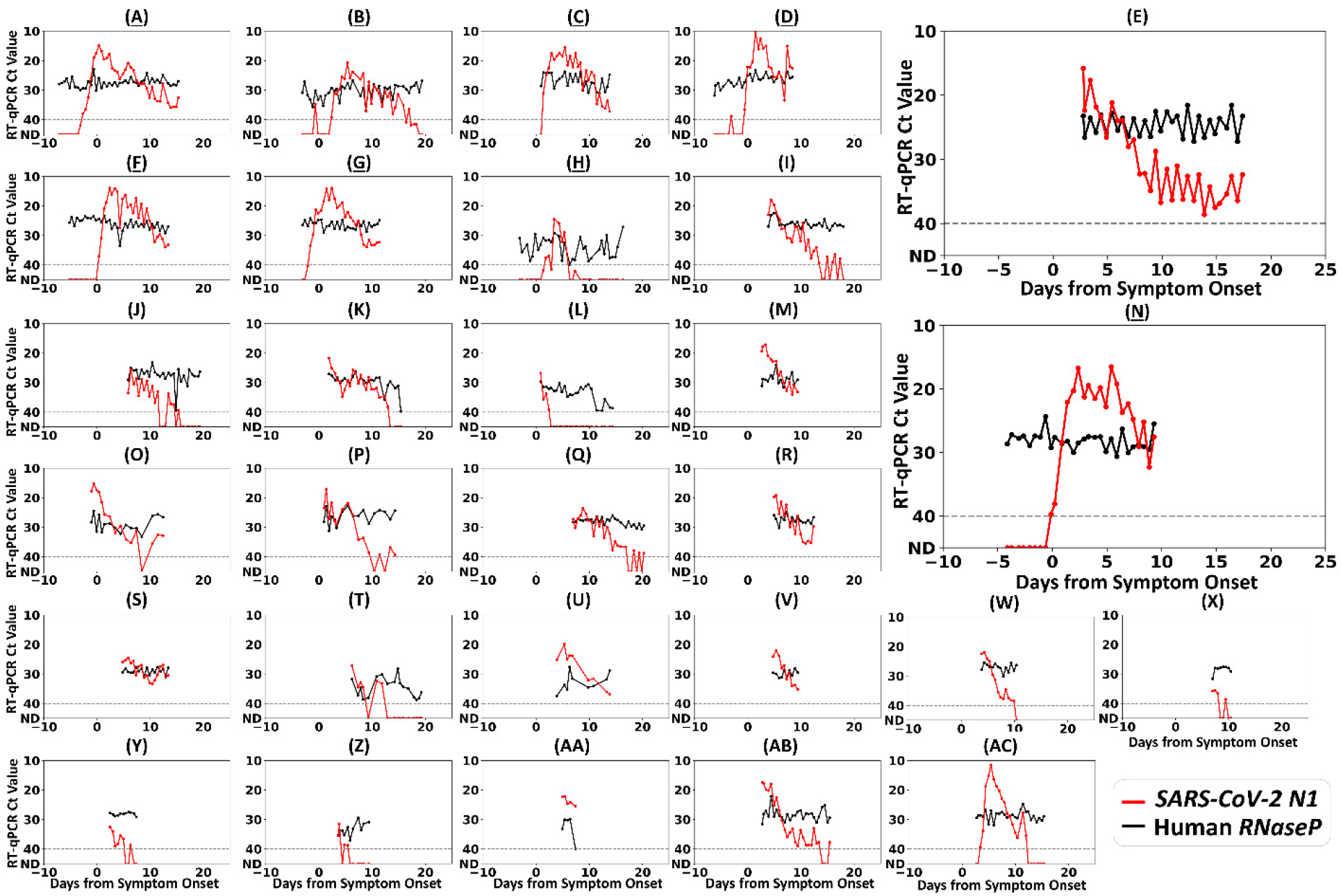
Individual nasal-swab RT-PCR Ct measurements, for SARS-CoV-2 *N1* gene target and human *RNase P* control gene target. Each panel shows the measured *SARS-CoV-2 N1* Ct values (red), and human *RNase P* Ct values (black) for an individual participant, relative to symptom onset. Matching panel labels correspond to the same participant shown in Figure S2. Underlined panel labels indicate that the participant converted from SARS-CoV-2-negative to -positive while enrolled in the study. Grey dashed line indicates Ct threshold for positivity. ND indicates Not Detected.

**Figure S4.**
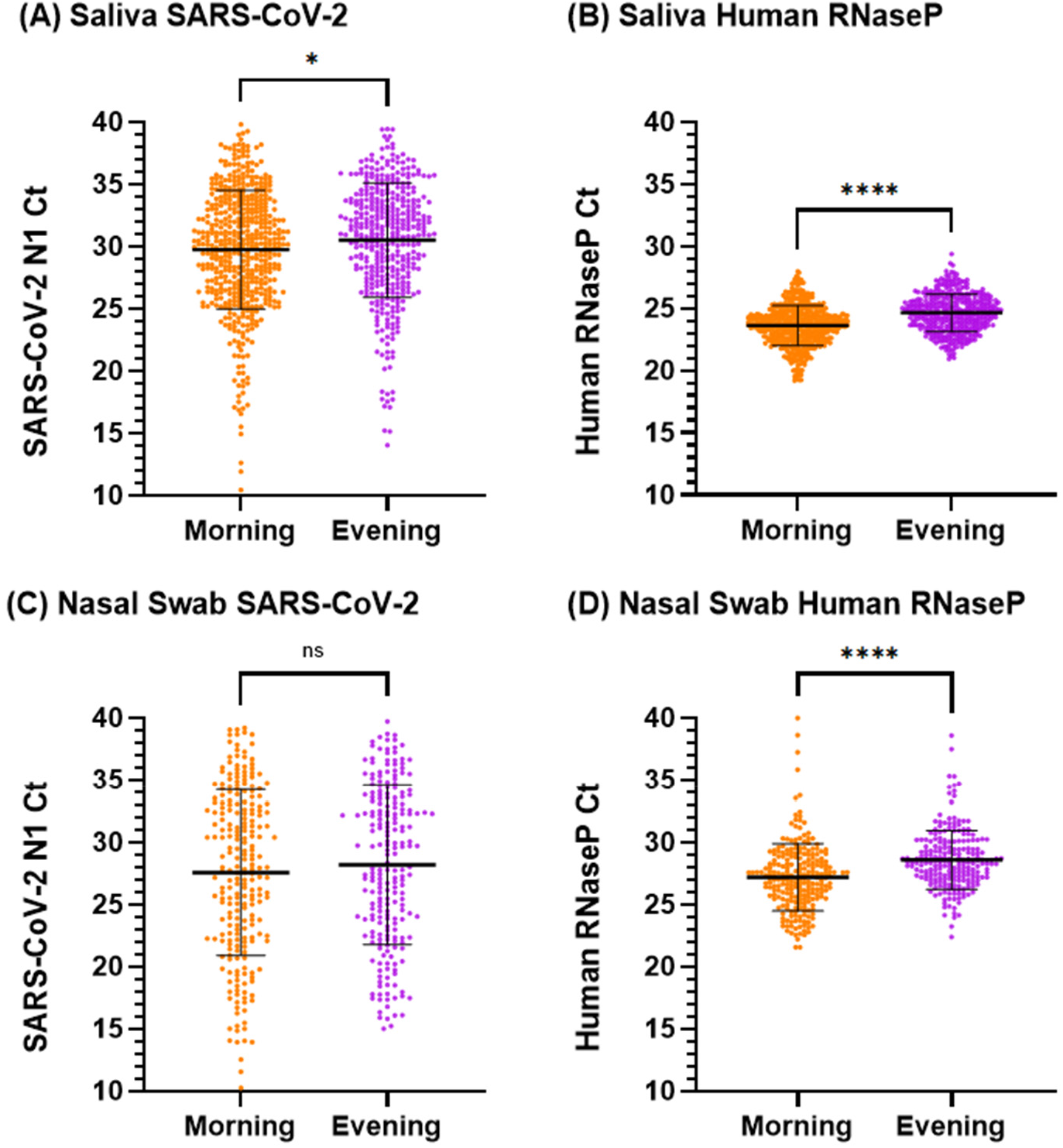
Aggregated SARS-CoV-2 *N1* and human *RNase P* Ct values grouped by samples collected in the morning and evening. **A)** Direct comparison of aggregated Ct values for *SARS-CoV-2 N1* gene target, measured from all SARS-CoV-2 positive saliva samples from all participants, by either morning or evening collection time **B)** Direct comparison of aggregated Ct values for human *RNase P* target from all SARS-CoV-2 positive saliva samples from all participants, by either morning or evening collection time **C)** Direct comparison of aggregated Ct values for *SARS-CoV-2 N1* gene, measured from all SARS-CoV-2 positive nasal-swab samples from all participants, by either morning or evening collection time **D)** Direct comparison of aggregated Ct values for human *RNase P* target from all SARS-CoV-2 positive nasal-swab samples from all participants, by either morning or evening collection time. Samples with morning collection times are shown as orange points, while evening are shown as purple points. Black lines indicate mean Ct value, with error bars representing standard deviation. Statistical comparison of Ct values for groups performed by unpaired t-test without correction: ns indicates nonsignificant difference, * indicates *P* <0.05, **** indicates *P* <0.001.

## AUTHOR CONTRIBUTIONS (listed alphabetically by last name)

RA: collaborated with AVW in creating digital participant symptom surveys; assisted with data quality control/curation with JJ, NWS, NS; created current laboratory information management system (LIMS) for specimen logging and tracking. Creation of iOS application for sample logging/tracking. Configured an SQL database for data storage. Created an Apache server and websites to view study data. Configured FTPS server to catalog PCR data. Wrote a Python package to access study data. Worked with ESS, AVW, AER to implement logic that prioritized specimen extraction order.

JTB: Created initial specimen tracking database to aid in specimen logging and tracking. Maintenance of database and implementation of corrections.

AMC: Received, performed QC on, and logged specimens. Performed nucleic acid extractions and RT-qPCR. Aliquoting and preparing study reagents as needed. Performed preliminary experiments to assess RNA stability in our saliva and nasal swab samples. Summarizing daily RT-qPCR data of participant time courses to inform participant keep/drop decisions. Provided feedback on early figure drafts.

MMC: Collaborated with AVW, MF, NS, YG, RFI, on study design and recruitment strategies. Co-wrote initial IRB protocol and informed consent with AVW and NS; assisted in the writing of the enrollment questionnaire; developed laboratory sample processing workflow for saliva with AVW and AER; performed sample processing on subset of samples; funding acquisition; collaborated with AER to write data processing/visualization code for observing household transmission events for active study participants. Contributor to the design of the calibration curve for saliva LOD experiments. Performed specimen logging and QC.

MF: Co-investigator; collaborated with AVW, MMC, NS, YG, RFI on study design and recruitment strategies; provided guidance and expertise on SARS-CoV-2 epidemiology and local trends.

YG: Co-investigator; collaborated with AVW, MMC, NS, MF, RFI on study design and recruitment strategies; provided guidance and expertise on SARS-CoV-2 epidemiology and local trends.

RFI: Co-investigator; collaborated with AVW, MMC, NS, MF, YYG on study design and recruitment strategies; provided leadership, technical guidance, oversight, and was responsible for obtaining funding for the study.

JJ: Researched epidemiological survey structures, performed epidemiological literature review with MMC and AVW, and co-wrote enrollment questionnaire with NS and AVW. Major contributor to curation of participant symptom data. Provided quality control of participant data with RA, NS, NWS.

MKP: Performed specimen logging and QC, RNA extractions, RT-qPCR, data processing. Performed data acquisition and analysis for and made Figure S2 with AVW. Prepared participant sample collection materials and helped with supplies acquisition. Assisted in literature analysis with ESS, RA, AVW. Performed data analysis and prepared Fig. 1B, 1D with AVW. Performed data analysis and prepared Fig. 2B, 2D with AVW. Verified the underlying data with AVW and NS. Outlined and wrote manuscript with AVW.

JAR: Lead study coordinator; collaborated with NS, AVW, NWS, and RFI on recruitment strategies, translated study materials into Spanish, co-wrote informational sheets with AVW and NS; created instructional videos for participants; enrolled and maintained study participants with NS and NWS.

AER: Developed laboratory swab sample processing workflow with ESS. Optimized extraction protocols working with vendor scientists. Created budgets and managed, planned, and purchased reagents and supplies; developed and validated method for RT-qPCR analysis for saliva and nasal-swab samples with MMC, and AVW. Performed specimen logging and QC, RNA extractions, RT-qPCR; Design of saliva calibration curve experiment. Managing logistics for the expansion of the BSL-2+ lab space with ESS. Provided feedback on earlier manuscript revision and provided a few references.

ESS: Coordinated laboratory team schedules and division of lab work. Performed initial nasal-swab workflow validation experiments with AER. Major contributor to workflow validation, methods, biosafety SOPs. Developed/implemented system for sample archiving. Performed specimen logging and QC, RNA extractions, RT-qPCR, and data processing. Managing logistics for the expansion of the BSL-2+ lab space with AER and biohazardous waste pickups.

NS: Study administrator; collaborated with AVW, MMC, RFI, YG, MF on initial study design and recruitment strategies; co-wrote IRB protocol and informed consent with AVW and MMC; co-wrote enrollment questionnaire with AVW and JJ; co-wrote participant informational sheets with AVW and JAR; enrolled and maintained study participants with JAR and NWS; study-data quality control, curation and archiving with RA, JJ, and NWS; reagents and supplies acquisition; assembled Table 1 with AVW; managed citations and reference library; verified the underlying data with MKP and AVW; edited the manuscript.

NWS: Study coordinator; collaborated with NS, AW, JAR, and RFI on recruitment strategies; enrolled and maintained study participants with NS and JAR; study-data quality control, curation and archiving with RA, JJ, and NS.

CT: Coordinated the recruitment efforts at PPHD with case investigators and contact tracers; provided guidance and expertise on SARS-CoV-2 epidemiology and local trends

AVW: Collaborated with MMC, NS, RFI, YG, MF on initial study design and recruitment strategies; co-wrote IRB protocol and informed consent with MMC and NS; co-wrote enrollment questionnaire with NS and JJ; co-wrote participant informational sheets with NS and JAR and digital survey; developed and validated methods for saliva and nasal-swab sample collection; developed and validated methods for RT-qPCR analysis for saliva and swab samples with AER, ESS, MMC; reagents and supplies acquisition; funding acquisition; developed laboratory sample processing workflow with AER, ESS, and MMC; performed specimen logging and QC, nucleic acid extraction, RT-qPCR; assembled Table 1 with NS; analyzed viral load timeseries data to visualize trends (Fig. 1A, Fig. 1C, Fig. 2A, Fig. 2C); assisted MKP in the preparation of Fig. 1B, 1D; analyzed viral load data to generate Fig 3; analyzed sample collection data for Fig S1; generated longitudinal RT-qPCR plot array for Fig S2, Fig S3; analyzed data for Fig S4. Verified the underlying data with MKP and NS. Outlined and wrote manuscript with MKP.

## REFERENCES

1. A. Wadhwa, Fisher, K.A., Silver, R., Koh, M., Arons, M.M., Miller, D.A., McIntyre, A.F., Vuong, J.T., Kim, K., Takamiya, M., Binder, A.M., Tate, J.E., Armstrong, P.A., Black, S.R., Mennella, C.C., Levin, R., Gubser, J., Jones, B., Welbel, S.F., Moonan, P.K., Curran, K., Ghinai, I., Doshi, R. and Zawitz, C.J. 2021. “Identification of Presymptomatic and Asymptomatic Cases Using Cohort-Based Testing Approaches at a Large Correctional Facility—Chicago, Illinois, USA, May 2020.” Clinical Infectious Diseases. 72: e128–e135.

2. Dora AV, Winnett A, Jatt LP and et al. 2020. “Universal and Serial Laboratory Testing for SARS-CoV-2 at a Long-Term Care Skilled Nursing Facility for Veterans — Los Angeles, California, 2020.” MMWR Morb Mortal Wkly Rep. 69: 651–655.

3. J. Lai, German, J., Hong, F., Tai, S.H.S., McPhaul, K.M., Milton, D.K. and for the University of Maryland Stop, C.R.G. 2022. “Comparison of Saliva and Mid-Turbinate Swabs for Detection of COVID-19.” medRxiv. 2021.2012.2001.21267147.

4. A.L. Wyllie, Fournier, J., Casanovas-Massana, A., Campbell, M., Tokuyama, M., Vijayakumar, P., Warren, J.L., Geng, B., Muenker, M.C., Moore, A.J., Vogels, C.B.F., Petrone, M.E., Ott, I.M., Lu, P., Venkataraman, A., Lu-Culligan, A., Klein, J., Earnest, R., Simonov, M., Datta, R., Handoko, R., Naushad, N., Sewanan, L.R., Valdez, J., White, E.B., Lapidus, S., Kalinich, C.C., Jiang, X., Kim, D.J., Kudo, E., Linehan, M., Mao, T., Moriyama, M., Oh, J.E., Park, A., Silva, J., Song, E., Takahashi, T., Taura, M., Weizman, O.-E., Wong, P., Yang, Y., Bermejo, S., Odio, C.D., Omer, S.B., Dela Cruz, C.S., Farhadian, S., Martinello, R.A., Iwasaki, A., Grubaugh, N.D. and Ko, A.I. 2020. “Saliva or Nasopharyngeal Swab Specimens for Detection of SARS-CoV-2.” New England Journal of Medicine. 383: 1283–1286.

5. R. Ke, Martinez, P.P., Smith, R.L., Gibson, L.L., Mirza, A., Conte, M., Gallagher, N., Luo, C.H., Jarrett, J., Conte, A., Liu, T., Farjo, M., Walden, K.K.O., Rendon, G., Fields, C.J., Wang, L., Fredrickson, R., Edmonson, D.C., Baughman, M.E., Chiu, K.K., Choi, H., Scardina, K.R., Bradley, S., Gloss, S.L., Reinhart, C., Yedetore, J., Quicksall, J., Owens, A.N., Broach, J., Barton, B., Lazar, P., Heetderks, W.J., Robinson, M.L., Mostafa, H.H., Manabe, Y.C., Pekosz, A., McManus, D.D. and Brooke, C.B. 2021. “Daily sampling of early SARS-CoV-2 infection reveals substantial heterogeneity in infectiousness.” medRxiv. 2021.2007.2012.21260208.

6. R. Wölfel, Corman, V.M., Guggemos, W., Seilmaier, M., Zange, S., Müller, M.A., Niemeyer, D., Jones, T.C., Vollmar, P., Rothe, C., Hoelscher, M., Bleicker, T., Brünink, S., Schneider, J., Ehmann, R., Zwirglmaier, K., Drosten, C. and Wendtner, C. 2020. “Virological assessment of hospitalized patients with COVID-2019.” Nature. 581: 465–469.

7. R.L. Smith, Gibson, L.L., Martinez, P.P., Ke, R., Mirza, A., Conte, M., Gallagher, N., Conte, A., Wang, L., Fredrickson, R., Edmonson, D.C., Baughman, M.E., Chiu, K.K., Choi, H., Jensen, T.W., Scardina, K.R., Bradley, S., Gloss, S.L., Reinhart, C., Yedetore, J., Owens, A.N., Broach, J., Barton, B., Lazar, P., Henness, D., Young, T., Dunnett, A., Robinson, M.L., Mostafa, H.H., Pekosz, A., Manabe, Y.C., Heetderks, W.J., McManus, D.D. and Brooke, C.B. 2021. “Longitudinal assessment of diagnostic test performance over the course of acute SARS-CoV-2 infection.” medRxiv.

8. A. Singanayagam, Hakki, S., Dunning, J., Madon, K.J., Crone, M.A., Koycheva, A., Derqui-Fernandez, N., Barnett, J.L., Whitfield, M.G., Varro, R., Charlett, A., Kundu, R., Fenn, J., Cutajar, J., Quinn, V., Conibear, E., Barclay, W., Freemont, P.S., Taylor, G.P., Ahmad, S., Zambon, M., Ferguson, N.M., Lalvani, A., Badhan, A., Dustan, S., Tejpal, C., Ketkar, A.V., Narean, J.S., Hammett, S., McDermott, E., Pillay, T., Houston, H., Luca, C., Samuel, J., Bremang, S., Evetts, S., Poh, J., Anderson, C., Jackson, D., Miah, S., Ellis, J. and Lackenby, A. “Community transmission and viral load kinetics of the SARS-CoV-2 delta (B.1.617.2) variant in vaccinated and unvaccinated individuals in the UK: a prospective, longitudinal, cohort study.” The Lancet Infectious Diseases.

9. S. Savela Emily, Viloria Winnett, A., Romano Anna, E., Porter Michael, K., Shelby, N., Akana, R., Ji, J., Cooper Matthew, M., Schlenker Noah, W., Reyes Jessica, A., Carter Alyssa, M., Barlow Jacob, T., Tognazzini, C., Feaster, M., Goh, Y.-Y. and Ismagilov Rustem, F. “Quantitative SARS-CoV-2 viral-load curves in paired saliva and nasal swabs inform appropriate respiratory sampling site and analytical test sensitivity required for earliest viral detection.” Journal of Clinical Microbiology. 0: JCM.01785-01721.

10. S.M. Kissler, Fauver, J.R., Mack, C., Tai, C.G., Breban, M.I., Watkins, A.E., Samant, R.M., Anderson, D.J., Metti, J., Khullar, G., Baits, R., MacKay, M., Salgado, D., Baker, T., Dudley, J.T., Mason, C.E., Ho, D.D., Grubaugh, N.D. and Grad, Y.H. 2021. “Viral Dynamics of SARS-CoV-2 Variants in Vaccinated and Unvaccinated Persons.” New England Journal of Medicine. 385: 2489–2491.

11. S.M. Kissler, Fauver, J.R., Mack, C., Olesen, S.W., Tai, C., Shiue, K.Y., Kalinich, C.C., Jednak, S., Ott, I.M., Vogels, C.B.F., Wohlgemuth, J., Weisberger, J., DiFiori, J., Anderson, D.J., Mancell, J., Ho, D.D., Grubaugh, N.D. and Grad, Y.H. 2021. “Viral dynamics of acute SARS-CoV-2 infection and applications to diagnostic and public health strategies.” PLOS Biology. 19: e3001333.

12. FDA. 2020. “SARS-CoV-2 Reference Panel Comparative Data.” https://www.fda.gov/medical-devices/coronavirus-covid-19-and-medical-devices/sars-cov-2-reference-panel-comparative-data

13. L.E. Brümmer, Katzenschlager, S., Gaeddert, M., Erdmann, C., Schmitz, S., Bota, M., Grilli, M., Larmann, J., Weigand, M.A., Pollock, N.R., Macé, A., Carmona, S., Ongarello, S., Sacks, J.A. and Denkinger, C.M. 2021. “Accuracy of novel antigen rapid diagnostics for SARS-CoV-2: A living systematic review and meta-analysis.” PLOS Medicine. 18: e1003735.

14. WHO. 2020. “Global partnership to make available 120 million affordable, quality COVID-19 rapid tests for low-and middle-income countries.” https://www.who.int/news/item/28-09-2020-global-partnership-to-make-available-120-million-affordable-quality-covid-19-rapid-tests-for-low--and-middle-income-countries

15. Health Affairs. 2021. “Access To COVID-19 Testing In Low-And Middle-Income Countries Is Still Critical To Achieving Health Equity.” https://www.healthaffairs.org/do/10.1377/forefront.20211026.483412/full/

16. PATH. 2022. “Global Availability of COVID-19 Diagnostic Tests.” https://www.path.org/programs/diagnostics/covid-dashboard-global-availability-covid-19-diagnostic-tests/

17. H. Borrmann, McKeating, J.A. and Zhuang, X. 2020. “The Circadian Clock and Viral Infections.” Journal of Biological Rhythms. 36: 9–22.

18. A.B. Diallo, Coiffard, B., Leone, M., Mezouar, S. and Mege, J.-L. 2020. “For Whom the Clock Ticks: Clinical Chronobiology for Infectious Diseases.” Frontiers in Immunology. 11:

19. A. Winnett, Cooper, M.M., Shelby, N., Romano, A.E., Reyes, J.A., Ji, J., Porter, M.K., Savela, E.S., Barlow, J.T., Akana, R., Tognazzini, C., Feaster, M., Goh, Y.-Y. and Ismagilov, R.F. 2020. “SARS-CoV-2 Viral Load in Saliva Rises Gradually and to Moderate Levels in Some Humans.” medRxiv. 2020.2012.2009.20239467.

20. C.-G. Huang, Lee, K.-M., Hsiao, M.-J., Yang, S.-L., Huang, P.-N., Gong, Y.-N., Hsieh, T.-H., Huang, P.-W., Lin, Y.-J., Liu, Y.-C., Tsao, K.-C. and Shih, S.-R. 2020. “Culture-Based Virus Isolation To Evaluate Potential Infectivity of Clinical Specimens Tested for COVID-19.” Journal of clinical microbiology. 58: e01068–01020.

21. P. Virtanen, et al. 2020. “SciPy 1.0: fundamental algorithms for scientific computing in Python.” Nature Methods. 17: 261–272.

22. M. Liu, Li, Q., Zhou, J., Ai, W., Zheng, X., Zeng, J., Liu, Y., Xiang, X., Guo, R., Li, X., Wu, X., Xu, H., Jiang, L., Zhang, H., Chen, J., Tian, L., Luo, J. and Luo, C. 2020. “Value of swab types and collection time on SARS-COV-2 detection using RT-PCR assay.” Journal of Virological Methods. 286: 113974.

23. D.L.-L. Hung, Li, X., Chiu, K.H.-Y., Yip, C.C.-Y., To, K.K.-W., Chan, J.F.-W., Sridhar, S., Chung, T.W.-H., Lung, K.-C., Liu, R.W.-T., Kwan, G.S.-W., Hung, I.F.-N., Cheng, V.C.-C. and Yuen, K.-Y. 2020. “Early-Morning vs Spot Posterior Oropharyngeal Saliva for Diagnosis of SARS-CoV-2 Infection: Implication of Timing of Specimen Collection for Community-Wide Screening.” Open Forum Infectious Diseases. 7: ofaa210.

24. M. Rao, Rashid, F.A., Sabri, F.S.A.H., Jamil, N.N., Zain, R., Hashim, R., Amran, F., Kok, H.T., Samad, M.A.A. and Ahmad, N. 2021. “Comparing Nasopharyngeal Swab and Early Morning Saliva for the Identification of Severe Acute Respiratory Syndrome Coronavirus 2 (SARS-CoV-2).” Clinical Infectious Diseases. 72: e352–e356.

25. A. Bivins, North, D., Wu, Z., Shaffer, M., Ahmed, W. and Bibby, K. 2021. “Within-and between-Day Variability of SARS-CoV-2 RNA in Municipal Wastewater during Periods of Varying COVID-19 Prevalence and Positivity.” ACS ES&T Water. 1: 2097–2108.

26. FDA. 2019. “Home Use Tests: Pregnancy.” https://www.fda.gov/medical-devices/home-use-tests/pregnancy

27. I.F. Uludag, Tiftikcioglu, B.I. and Ertekin, C. 2016. “Spontaneous Swallowing during All-Night Sleep in Patients with Parkinson Disease in Comparison with Healthy Control Subjects.” Sleep. 39: 847–854.

28. C. Dawes. 1972. “Circadian rhythms in human salivary flow rate and composition.” J Physiol. 220: 529–545.

29. G. Mazzoccoli, Vinciguerra, M., Carbone, A. and Relógio, A. 2020. “The Circadian Clock, the Immune System, and Viral Infections: The Intricate Relationship Between Biological Time and Host-Virus Interaction.” Pathogens (Basel, Switzerland). 9: 83.

30. J.E. Niemeyer. 2017. “Viruses and circadian rhythms.” Lab Animal. 46: 7–7.

31. A.B. Diallo, Gay, L., Coiffard, B., Leone, M., Mezouar, S. and Mege, J.-L. 2021. “Daytime variation in SARS-CoV-2 infection and cytokine production.” Microbial Pathogenesis. 158: 105067.

32. S. Ray and Reddy, A.B. 2020. “COVID-19 management in light of the circadian clock.” Nature Reviews Molecular Cell Biology. 21: 494–495.

33. M. Meira e Cruz, Miyazawa, M. and Gozal, D. 2020. “Putative contributions of circadian clock and sleep in the context of SARS-CoV-2 infection.” European Respiratory Journal. 55: 2001023.

34. X. Zhuang, et al. 2021. “The circadian clock component BMAL1 regulates SARS-CoV-2 entry and replication in lung epithelial cells.” iScience. 24: 103144.

35. E.N. Gallichotte, Quicke, K.M., Sexton, N.R., Fitzmeyer, E., Young, M.C., Janich, A.J., Dobos, K., Pabilonia, K.L., Gahm, G., Carlton, E.J., Ebel, G.D. and Ehrhart, N. 2020. “Longitudinal Surveillance for SARS-CoV-2 Among Staff in Six Colorado Long-Term Care Facilities: Epidemiologic, Virologic and Sequence Analysis.” medRxiv : the preprint server for health sciences. 2020.2006.2008.20125989.

36. R. Perera, Tso, E., Tsang, O., Tsang, D., Fung, K., Leung, Y. and et al. 2020. “SARS-CoV-2 Virus Culture and Subgenomic RNA for Respiratory Specimens from Patients with Mild Coronavirus Disease.” Emerg Infect Dis. 26: 2701–2704.

37. I.N. Okeke and Ihekweazu, C. 2021. “The importance of molecular diagnostics for infectious diseases in low-resource settings.” Nature Reviews Microbiology. 19: 547–548.

38. A.-L. Sander, et al. “An Observational Laboratory-Based Assessment of SARS-CoV-2 Molecular Diagnostics in Benin, Western Africa.” mSphere. 6: e00979–00920.

39. The Lancet Global Health. 2021. “Essential diagnostics: mind the gap.” The Lancet Global Health. 9: e1474.

40. A. Terriau, Albertini, J., Montassier, E., Poirier, A. and Le Bastard, Q. 2021. “Estimating the impact of virus testing strategies on the COVID-19 case fatality rate using fixed-effects models.” Scientific Reports. 11: 21650.

